# Association of Interleukin-6 gene (*rs1800795*) variant with waist-hip-ratio and Insulin resistance: A cross sectional study in adult women

**DOI:** 10.1101/2024.05.24.24307852

**Authors:** Abhishek Gupta, Arun Kumar Singh, Priyanka Gupta, Komal Shah, Vani Gupta

## Abstract

Obesity is a major contributing factor to metabolic disorders and public health problems worldwide. The goal of the current study was to determine if the Interleukin-6 gene rs1800795 variant is linked to waist-hip-ratio (WHR) and insulin resistance (IR) in North Indian adult women. The WHO guidelines were followed by 37 women who had a WHR >0.85 and 35 women who had a WHR <0.85, out of the 72 women who took part in the study. Every adult woman underwent anthropometric measures. Fasting blood glucose, serum Insulin, lipid levels, and IL-6 levels were measured, and IR was computed. The RFLP technique was used to polymorphise the IL-6 gene (174 G>C) variant. The study group’s Insulin, HOMA-IR, BMI, lipid profile, TC/HDL ratio, HDL/LDL ratio, and serum IL-6 level were shown to differ significantly from the control. The percentage frequency of the IL-6 mutant genotype GC+CC (89.19%) and mutant allele C (75.68%) was higher in the study group (WHR >0.85). According to the findings, WHR, TC, TG, and HOMA-IR were substantially correlated (all p <0.05) with the mutant genotype GC+CC of the IL-6 174 G>C gene. This study indicates that the IL-6 rs1800795 gene (174 G>C) variant was substantially linked with metabolic risk variables, such as WHR and HOMA-IR.

## 1. Introduction

Globally, obesity is a significant contributing factor to metabolic diseases and issues with public health. It is characterized by poor lifestyle choices, high calorie fat foods, and minimal physical activity. Obesity is a risk factor for diabetes that is associated with insulin resistance (IR). Type II diabetes (T2), hypertension, and cardiovascular diseases (CVDs) may be triggered by this (*1*). It is an increase in the quantity of adipose tissue (AT), and the production of components by AT that makes some people more IR than others is one explanation for obesity-related IR. As both increase waist-hip-ratio (WHR) and IR are emerging risk factors associated with abdominal obesity and said to be more genetically predisposed for the same in Indians. All classifications including EGIR, WHO, and NCEP-ATP III 2001, recognize the importance of abdominal obesity (*2*). Conversely, the WHO definition suggests use the waist-hip ratio (WHR) in place of waist circumferences (WC), and the NCEP definition provides the loosest criteria for WC.

Increase in hormonal level, pro-inflammatory cytokine, and non-esterified fatty acid levels are released by AT in centrally obese individuals, and these factors may play a role in the emergence of IR. Interleukin-6 (IL-6) is a cytokine that affects the immune system and inflammation. Evidence for genetic control of human body weight and composition is well established. So far single gene mutations have only been identified as the cause of morbid obesity in young individuals in about 5% of the cases (*3-5*). Moreover, little is known about susceptibility genes contributing to modest obesity although knowledge is accumulating. There has been research on the relationship between many health issues and polymorphisms in the IL-6 location i.e., 174 G>C in the promoter region (*6-9*), including risk factors including IR and abdominal obesity in an estimated sample size study. However, no such data are available in the smaller study on IL-6 adipokines gene status and its correlation with abdominal obesity, which justify the effect of IL-6 variant on risk factors in an Indian adult woman. There is a conflicting body of research about the potential association between IR and the IL-6 174 G>C gene.

In the present study, we proposed that abdominal obesity is more likely among people with a WHR of greater than 0.85 who also had hypertriglyceridemia, lower HDL cholesterol, and higher IR. Abdominal obesity is more common among women who have polymorphism in the IL-6 promoter region. The primary aim of the study was to evaluate the IR status of obese (study group; WHR >0.85) women using the homeostatic model assessment index (HOMA-index) and compare them to non-obese (control group; WHR <0.85) adult women. Determine the impact of blood glucose, insulin, and lipid profile and IL-6 serum levels in women in the study group who are obese and in the control group who are non-obese. Moreover, to ascertain the IL-6 174 G>C gene’s frequency distribution pattern and correlation with anthropometric variables and metabolic risk factors in adult women who are obese and non-obese.

## 2 Material and Methods

### 2.1 Study population

A total of 72 women (37 study group women who had a WHR >0.85 and 35 control group women who had a WHR <0.85) between the ages of 20 and 40 were enrolled from the Department of Medicine, KGMU, for this shorter-term (one year) case control study. The study investigation was carried out in the Department of Physiology at King George’s Medical University (KGMU), Lucknow, Uttar Pradesh, India. Both the waist-hip ratio (WHR) and body mass index (BMI), which are indicators of abdominal obesity, can be used to quantify obesity. According to WHO standards for predicting obesity (*10-11*), women with a WHR <0.85 were classed as non-obese (study) in this study, whereas those with a WHR >0.85 were designated obese (control).

A questionnaire was used to collect data on menstruation history, diet, family history, and medical conditions. Each woman participating in the study was without any kind of metabolic, endocrine, inflammatory, cardiac, and respiratory disorders. All the women were also neither an alcoholic or diabetic. The study excluded women who were nursing, pregnant, or experiencing any type of obstetrical or gynecological issue, as well as those who were taking medication. The study was approved by the ethical committee of KGMU, Lucknow. Written informed consent was obtained from all the participants women.

### 2.2 Blood collection and biochemical analysis

A total of 6 ml of venous blood was taken in the morning following an overnight fast from each adult women participated in the study. Plasma and serum were extracted from each blood samples which were used in the determination of biochemical, blood glucose and lipid profiles. Remaining of that were taken and placed in EDTA vials to extract DNA using a commercial kit (Qiagen, USA). The GOD-POD method was used to estimate the plasma glucose (Randox Laboratories Ltd., Antrim, UK), lipid profile by enzymatic method (Randox Laboratories Ltd., Antrim, UK), and serum insulin was assessed by immuno-radiometric assay method (Immunotech Radiova, Prague). Serum IL-6 level was determined by a sandwich ELISA method (Diaclone, France). By using the HOMA-Index, IR was calculated using the equation: HOMA Index = [fasting Insulin (μU/I) x fasting glucose (mmol/l)/22.5] (*12*). The HOMA-index 3.6 was used to make a laboratory diagnosis of IR in people who did not meet the clinical and biological criteria for the condition.

### 2.3 IL-6 174 G>C promoter gene analysis by PCR

DNA was extracted from cellular blood components by the salting out method. Genotyping procedure for the detection of the IL-6 174 G>C gene was examined (*13*). All mutant and heterozygous samples of women were re-genotyped in order to increase the genotyping quality and validation, and only the results for reproducible and error-free samples were documented. IL-6 174 G>C gene was detected by PCR on the Thermo Cycler instrument (Bio-Rad Inc. Hercules, CA, USA) with subsequent restriction analysis of PCR products (RFLP). SfaN1 restricted fragment were visualized using a 2.0% agarose gel stained with ethidium bromide.

### 2.4 Statistical analysis

All data was statistically analyzed by using ‘unpaired student t test’ and multiple logistic regression and Fisher-exact test. Where appropriate, χ ^2^ analysis was performed. Odds ratios and 95% confidence interval are given accordingly. Data were summarized as means ± standard deviation (SD). P values of <0.05 were considered statistically significant. For statistical analysis, we used commercially available software (GraphPad Prism 8.0 version).

## 3 Result

### 3.1 Anthropometric variables and biochemical profile in study and control group women

The anthropometric variables and biochemical profile of the adult women in the study (obese) and control group (non-obese) are displayed in **Table 1**.

**Table 1.** Anthropometric and biochemical profile in control and study group women.

| S. No. | Parameters | Control group<br>(WHR <0.85, n=35) | Study group<br>(WHR >0.85, n=37) | p value |
| --- | --- | --- | --- | --- |
| 1. | Age (years) | 27.77 ± 4.45 | 25.68 ± 5.58 | NS |
| 2. | WHR | 0.79 ± 0.04 | 0.89 ± 0.02 | *** |
| 3. | BMI (kg/m<sup>2</sup>) | 21.81 ± 3.11 | 25.46 ± 5.72 | * |
| 4. | Blood glucose (mg/dl) | 97.57 ± 18.29 | 96.38 ± 12.91 | NS |
| 5. | TC (mg/dl) | 155.26 ± 29.90 | 173.87 ± 37.24 | * |
| 6. | TG (mg/dl) | 126.47 ± 34.64 | 123.29 ± 34.98 | NS |
| 7. | HDL (mg/dl) | 51.16 ± 16.18 | 44.62 ± 8.34 | * |
| 8. | LDL (mg/dl) | 82.29 ± 30.83 | 107.70 ± 31.91 | * |
| 9. | VLDL (mg/dl) | 25.23 ± 6.84 | 24.37 ± 6.60 | NS |
| 10. | TC / HDL | 3.35 ± 1.41 | 4.04 ± 1.23 | * |
| 11. | HDL/ LDL | 0.76 ± 0.49 | 0.45 ± 0.16 | * |
| 12. | Insulin (µl) | 7.43 ± 3.57 | 15.44 ± 7.48 | *** |
| 13. | HOMA-IR | 1.80 ± 0.97 | 3.74 ± 1.77 | ** |
| 14. | IL-6 (pg/ml) | 3.30 ± 2.12 | 6.02 ± 3.62 | ** |

Values shown as mean ± SD. p value \*\*\*<0.0001, \*\*<0.01, & \*<0.05 (significant) and non-significant (NS)
WHR, waist-hip-ratio; BMI, body mass index; TC, total cholesterol; TG, triglyceride; HDL, high density lipoprotein; LDL, low density lipoprotein; VLDL, very low-density lipoprotein; HOMA-IR, homeostatic model assessment-insulin resistance; IL-6, Interleukin-6

In the study group women, there were varying degrees of changes to the following data: WHR (0.89 ± 0.02), blood glucose (96.38 ± 12.91), TG (123.29 ± 34.98), HDL/LDL ratio (0.45 ± 0.16), Insulin (15.44 ± 7.48), IR (HOMA-IR) (3.74 ± 1.77) and serum IL-6 level (6.02 ± 3.62). In contrast, in the control group women, there were varying degrees of changes to WHR (0.79 ± 0.04), blood glucose (97.57 ± 18.29), TG (126.47 ± 34.64), HDL/LDL ratio (0.76 ± 0.49), Insulin (7.43 ± 3.57), IR (HOMA-IR) (1.80 ± 0.97) and serum IL-6 level (3.30 ± 2.12). In the study group women, highly significant p values were discovered such as WHR, insulin, and IR (calculated HOMA-IR) (all p <0.0001) and serum IL-6 level (p<0.01). Additionally, significant p values (all p <0.05) were seen for BMI, TC, HDL, LDL, TC/HDL, and HDL/LDL in the obese group women **(Fig 1 & Fig 2)**.

**Figure 1.**
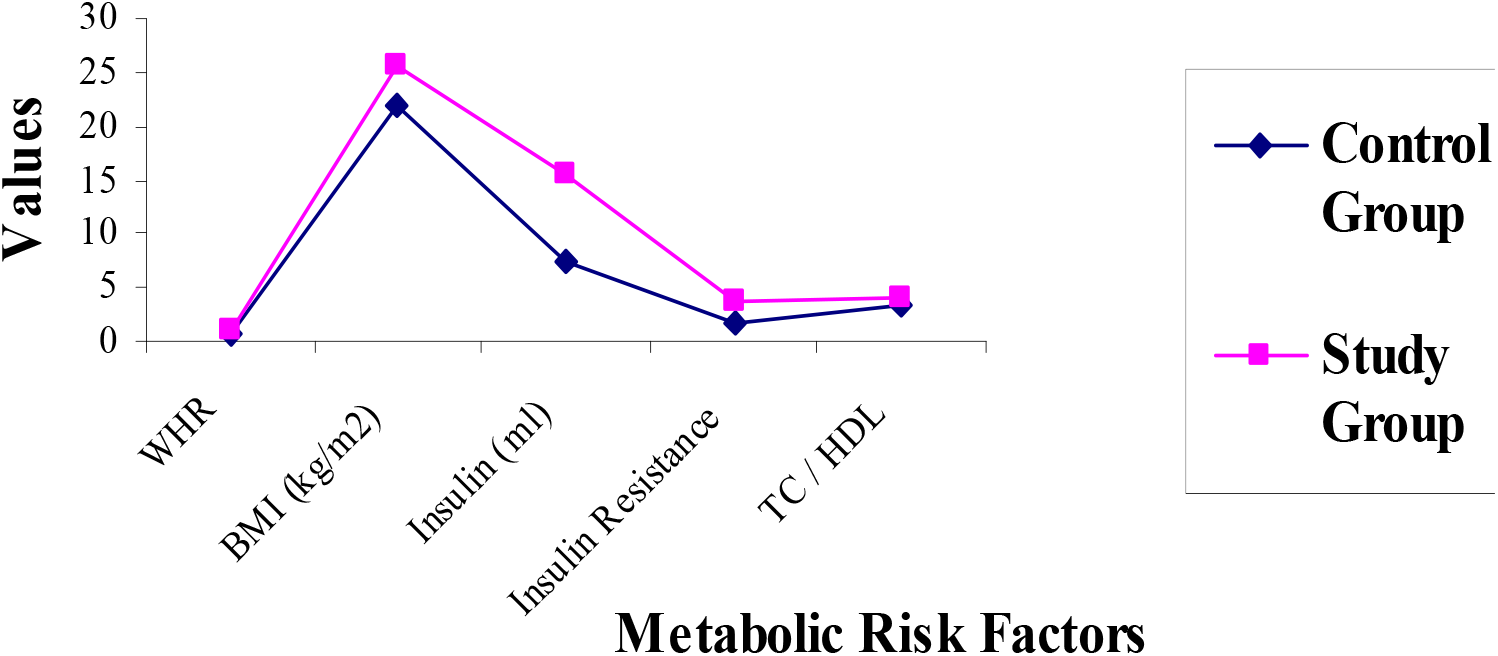
Metabolic risk factors (WHR, BMI, Insulin, IR and TC/HDL ratio) in study and control group women. WHR, waist-hip-ratio; BMI, body mass index; IR, insulin resistance; TC/HDL, total cholesterol/high-density lipoprotein

**Figure 2.**
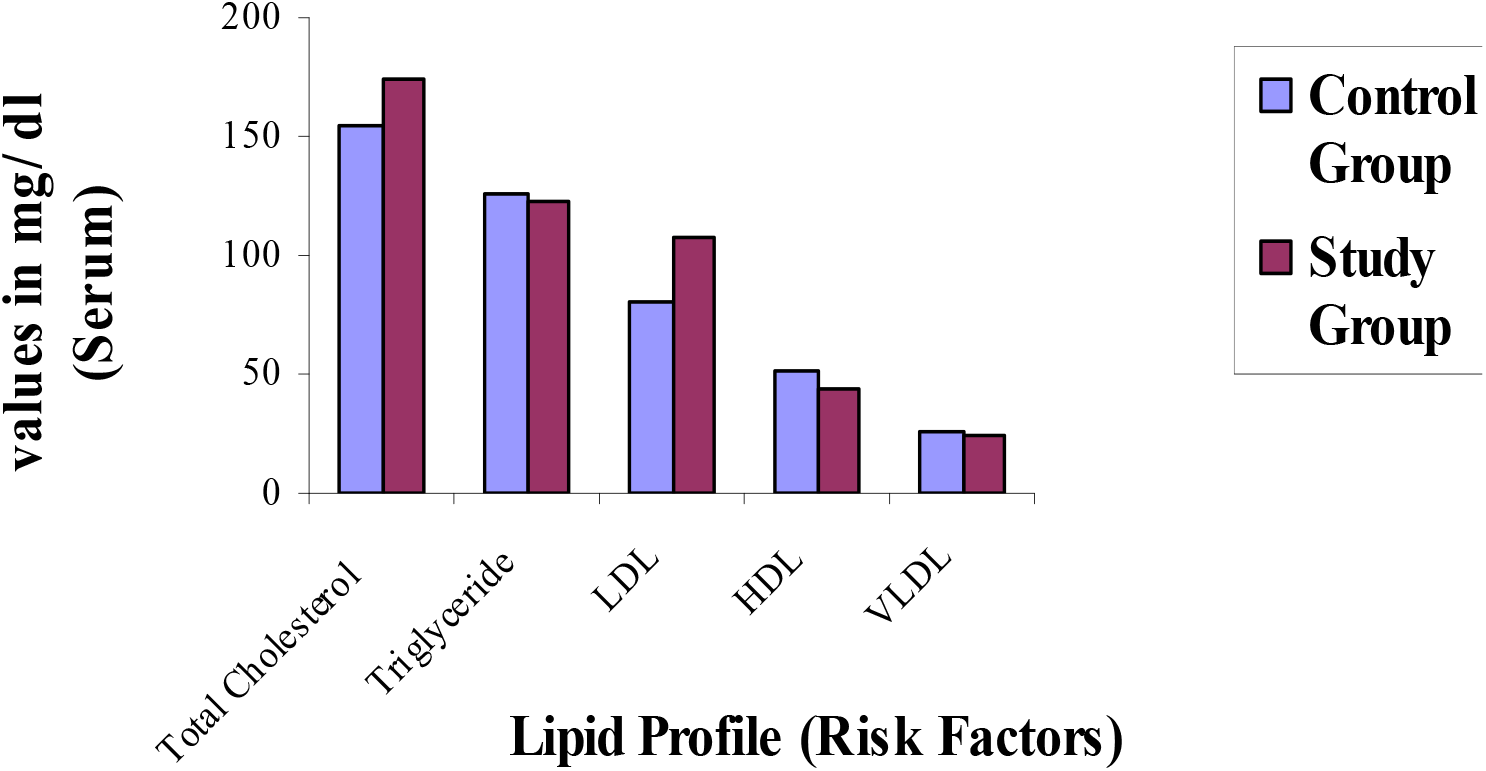
Lipid profile (TC, TG, LDL, HDL and VLDL) in study and control group women. TC, total cholesterol; TG, triglyceride; LDL, low-density lipoprotein; HDL, high density lipoprotein; VLDL, very low-density lipoprotein

### 3.2 Frequency distribution pattern of genotypes and alleles in IL-6 gene variant

On the other hand, allele and genotype frequencies of IL-6 174 G>C gene variant in study and control group are displayed in **Table 2**.

**Table 2.**
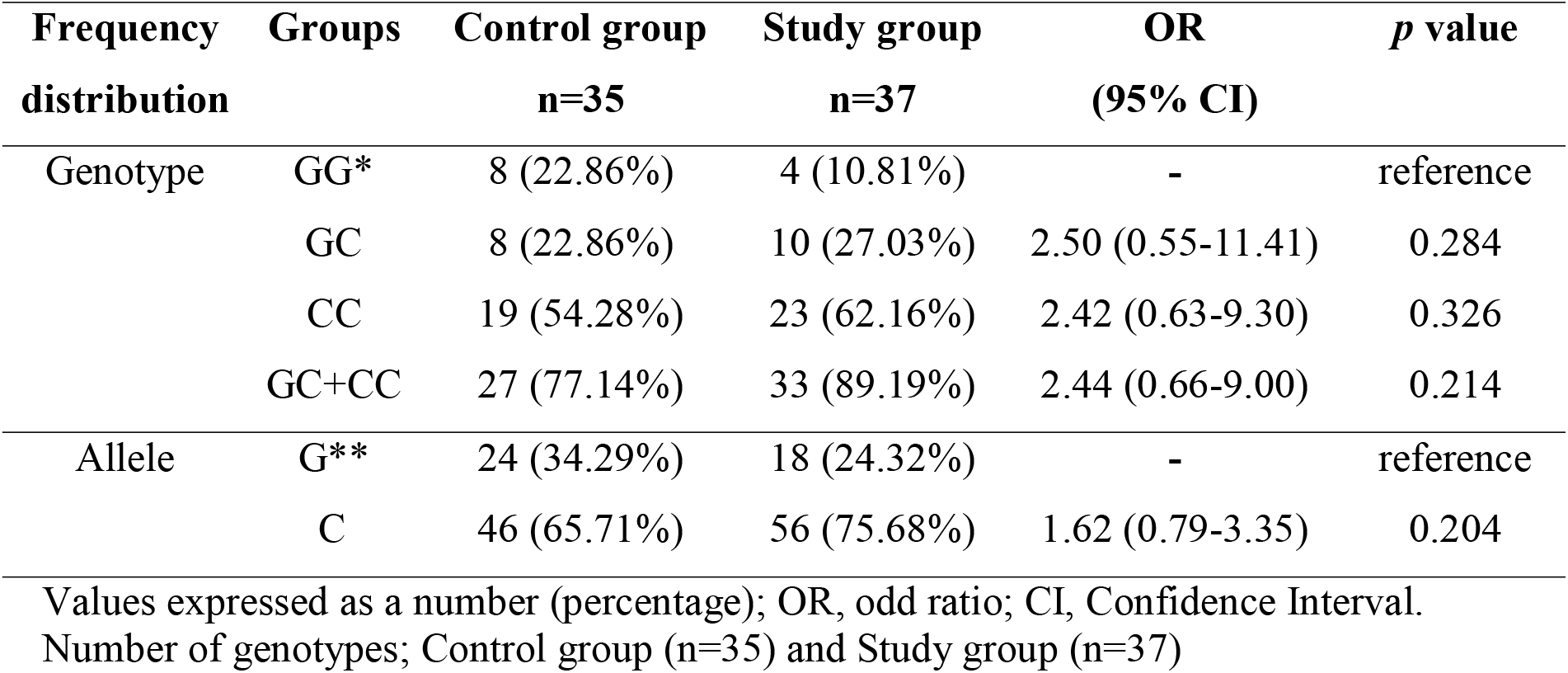
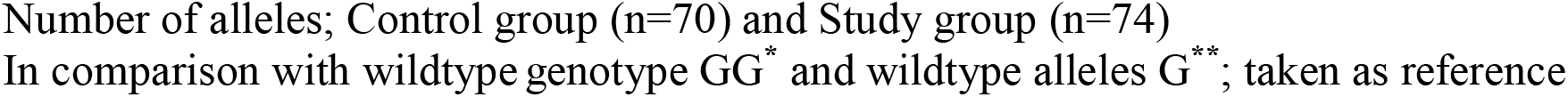
Allele and genotypes frequencies of IL-6 174 G>C gene polymorphism among control (WHR <0.85) and study (WHR >0.85) group women.

| Frequency distribution | Groups | Control group<br>n=35 | Study group<br>n=37 | OR<br>(95% CI) | p value |
| --- | --- | --- | --- | --- | --- |
| Genotype | GG* | 8 (22.86%) | 4 (10.81%) | - | reference |
|  | GC | 8 (22.86%) | 10 (27.03%) | 2.50 (0.55-11.41) | 0.284 |
|  | CC | 19 (54.28%) | 23 (62.16%) | 2.42 (0.63-9.30) | 0.326 |
|  | GC+CC | 27 (77.14%) | 33 (89.19%) | 2.44 (0.66-9.00) | 0.214 |
| Allele | G** | 24 (34.29%) | 18 (24.32%) | - | reference |
|  | C | 46 (65.71%) | 56 (75.68%) | 1.62 (0.79-3.35) | 0.204 |

Values expressed as a number (percentage); OR, odd ratio; CI, Confidence Interval. Number of genotypes; Control group (n=35) and Study group (n=37)
Number of alleles; Control group (n=70) and Study group (n=74) In comparison with wildtype genotype GG\* and wildtype alleles G\*\* ; taken as reference

The frequency for the wild type G allele was 24.32% and 34.29%, and for the mutant C allele was 75.68% and 65.71%, in study and control group, respectively. In the study group (WHR >0.85), there was a greater percentage frequency of the IL-6 mutant genotype GC+CC (89.19%) and mutant allele C (75.68%) than control group.

### 3.3 Association of IL-6 gene variant and metabolic risk factors

Women in the obese group with a WHR >0.85, the relationship between metabolic risk factors and the IL-6 gene variant are shown in **Table 3**.

**Table 3.**
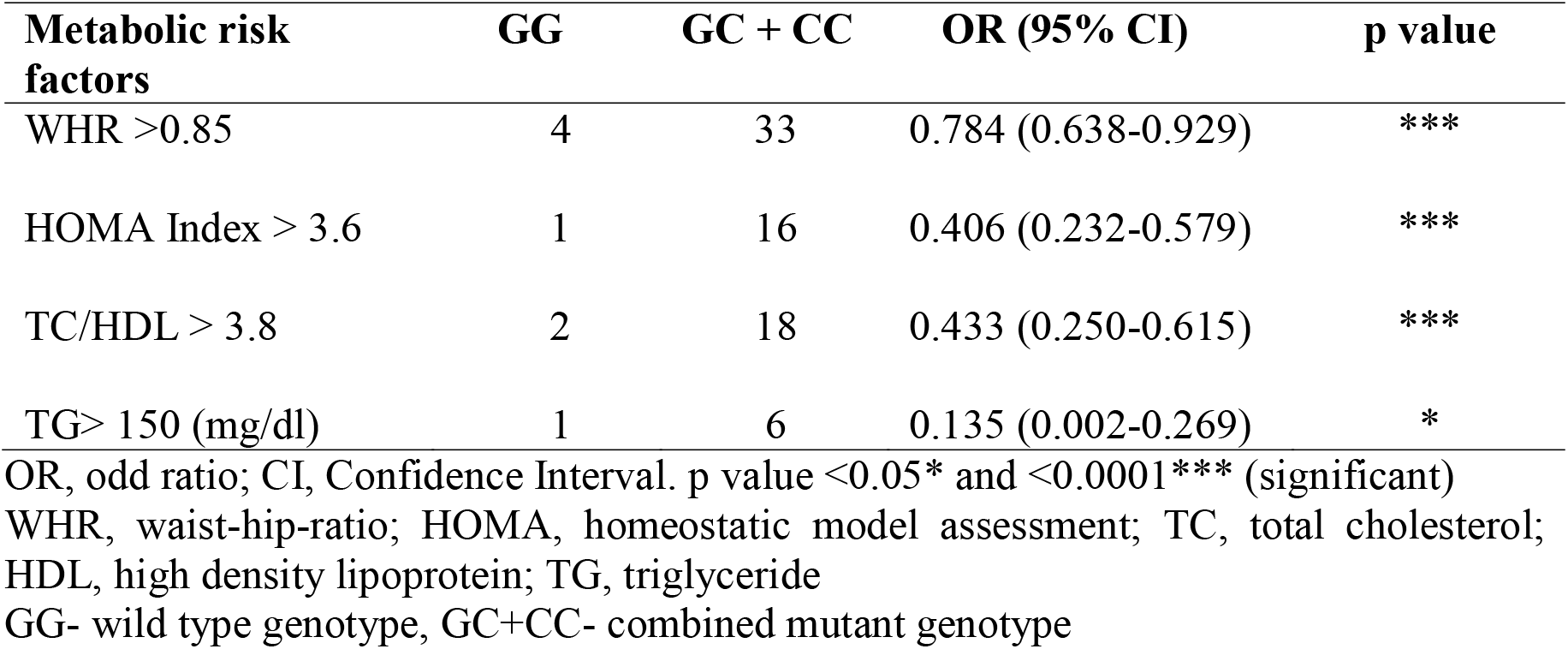
Association of IL-6 174 G>C gene and metabolic risk factors in study group women (WHR >0.85, n=37)

Significant association was observed between at least one mutant G allele of the IL-6 gene variant (174 G>C) and high WHR >0.85 (p=0.0001), HOMA-index >3.6 (p=0.0001), serum TC/HDL >3.8 (p=0.0001), and serum TG >150 mg/dl (p=0.047) were noted.

## 4 Discussion

The current study’s findings have a significant influence on creating a healthy environment. One of the most hazardous aspects of an unhealthy body is obesity. Individuals that suffer with this metabolic disorder don’t live their lives to the fullest. The current research indicates that adipokine synthesis and release occur primarily in human AT in obese individuals. It has been demonstrated that abdominal obesity raises the risk of metabolic diseases (*14,15*) including diabetes (*16*) as well as coronary heart disease (*17*). What causes the higher risk of fat storage in the abdominal area compared to general adiposity is yet unknown, though?

There are also notable variations in serum TC, LDL, HDL levels, HDL/LDL and TC/HDL, as well as in the levels of insulin, and IR in control and study women. This observation implies that one possible role for the linked variations may be in the development and cause of metabolic syndrome. The body’s actual fat deposition will be aided by the results of the HDL/LDL ratio. The WHR is a significant contributor to obesity and a key indicator of the disease’s progression. The accumulation of body fat that results in abdominal obesity is measured by taking measurements of the waist and hip circumferences. The correlation between rising adiposity and the development of IR implies that adipocyte products may play a significant role in IR.

Up to 30% of the total circulation concentration of IL-6 is secreted by AT (*18,19*), which may disrupt insulin signaling and cause IR in healthy individuals. Adipocyte-produced adipokine IL-6 is overexpressed in obese individuals with IR, and there is a link between the adipokine release. The results will encourage more research on the relationship between IR and IL-6 with WHR >0.85 in adult women, which implies that IR is the root cause of obesity. Additionally, obesity has been clearly linked to increased levels of circulating IL-6, body fat tissue, WC, and BMI. Higher levels of diabetes are correlated with higher levels of insulin sensitivity, and elevated levels of circulating IL-6 are linked to aberrant circulating lipid levels.

Polymorphism of adipokine gene at promoter region may suggest the alteration in transcription level, which may be led to metabolic changes and pathophysiology of IR. Several single-nucleotide polymorphisms (SNPs) and repeat polymorphisms have been described in the IL-6 gene region, but there are few known in the coding region or of intron/exon boundaries of the IL-6 gene (*20*). IL-6 gene transcription was found to be influenced by the C-174G polymorphism within the IL-6 promoter region (*13,21*). However, data about the effects of IL-6 gene variants on its IL-6 levels in humans are contradictory (*22,23*). The biallelic 174 IL-6 G>C SNP, which is located within the negative regulative domain of the IL-6 gene promoter, has been found to affect transcriptional regulation. It has been discovered that the IL-6 174 G>C gene affects transcription, leading to elevated IL-6 production in peripheral blood cells (*24,25*). The present study findings showed that combined mutant genotype GC+CC of the IL-6 gene was a high-risk genotype for the development of metabolic risk in comparison to their control counterpart. In line with the findings (*26*), we also observed a higher % frequency distribution of IL-6 gene in study women compared to control women. Additionally, we showed that women with mutant GC+CC genotypes had a significantly higher WHR than women with the wild GG genotype.

## 5 Conclusion

The study’s primary findings indicate that the obese group (WHR >0.85) had a significantly higher p values for metabolic factors such as WHR, TC, Insulin, HOMA-IR and serum IL-6 level. Thus, this study concluded that risk factors including WHR and HOMA-IR have strong correlation with the IL-6 rs1800795 gene (174G□>□C) variant. However, further research with larger sample sizes and addressing the identified limitations is warranted to validate and build upon these initial findings.

## Data Availability

All data produced in the present work are contained in the manuscript.

## Acknowledgements

The authors would like to thank the women participants who participated in this study. We extend our gratitude to the medical professionals and residents of King George’s Medical University (KGMU), Lucknow, India, for their kind assistance with this research.

## References

1. Petrie JR, Guzik TJ, Touyz RM. Diabetes, Hypertension, and cardiovascular disease: Clinical Insights and Vascular Mechanisms. Can J Cardiol. 2018; 34(5):575–584. doi: 10.1016/j.cjca.2017.12.005.

2. Gupta A, Gupta V. Metabolic syndrome: what are the risks for humans. Biosci Trends. 2010; 4(5):204–212.

3. Barsh GS, Farooqi IS, O’Rahilly S. Genetics of body-weight regulation. Nature 2000; 404:644–651.

4. Vaisse C, Clement K, Durand E, Hercberg S, Guy-Grand B, Froguel P. Melanocortin-4 receptor mutations are a frequent, heterogeneous cause of morbid obesity. J Clin Invest. 2000; 106:253–262.

5. Farooqi IS, Keogh JM, Yeo GS, Lank EJ, Cheetham T, O’Rahilly S. Clinical spectrum of obesity, mutations in the melanocortin 4 receptor gene. N Engl J Med 2003; 348:1085– 1095.

6. Gupta A, Gupta V, Shah K, Gupta V. Interleukin-6-174 G/C promoter gene polymorphism and polycystic ovary syndrome: A cross sectional investigation in adult women. Ann Clin Med Case Rep. 2024; 12(8):1–8. ACMCR-v12-2079.

7. Gupta A, Gupta P, Singh AK, Gupta V. Association of adipokines with insulin resistance and metabolic syndrome including obesity and diabetes. GHM Open. 2023; 3(1):7–19. doi: 10.35772/ghmo.2023.01004.

8. Shukla KK, Agnihotri S, Gupta A, Mahdi AA, Mohamed EA, Sankhwar SN, Sharma P. Significant association of TNFα and IL-6 gene with male infertility--an explorative study in Indian populations of Uttar Pradesh. Immunol Lett. 2013 Nov-Dec; 156(1-2):30–37. doi: 10.1016/j.imlet.2013.08.011.

9. Gupta A, Gupta V, Singh AK, Tiwari S, Agrawal S, Natu SM, Agrawal CG, Negi MP, Pant AB. Interleukin-6 G-174C gene polymorphism and serum resistin levels in North Indian women: potential risk of metabolic syndrome. Hum Exp Toxicol. 2011 Oct; 30(10):1445–1453. doi: 10.1177/0960327110393763. Epub 2010 Dec 22.

10. Geneva: World Health Organization; 2008. World Health Organization (WHO). Waist Circumference and Waist-Hip Ratio. Report of WHO Expert Consultation. 2008.

11. Nishida C, Ko GT, Kumanyika S. Body fat distribution and noncommunicable diseases in populations: overview of the 2008 WHO Expert Consultation on Waist Circumference and Waist-Hip Ratio. Eur J Clin Nutr. 2010; 64(1):2–5. doi: 10.1038/ejcn.2009.139. Epub 2009 Nov 25.

12. Matthews DR, Hosker JP, Rudenski AS, Naylor BA, Treacher DF, Turner RC. Homeostasis model assessment: insulin resistance and beta-cell function from fasting plasma glucose and insulin concentrations in man. Diabetologia. 1985; 28:412–419.

13. Fishman D, Faulds G, Jeffery R, Mohamed-Ali V, Yudkin JS, Humphries S, Woo P. The effect of novel polymorphisms in the interleukin-6 (IL-6) gene on IL-6 transcription and plasma IL-6 levels, and an association with systemic-onset juvenile chronic arthritis. J Clin Invest 1998; 102:1369–1376.

14. Von Eyben FE, Mouritsen E, Holm J, Montvilas P, Dimcevski G, Sucui G, Helleberg I, Kristensen L, von Eyben R. Intra-abdominal obesity and metabolic risk factors: a study of young adults. Int J Obes Relat Metab Disord. 2003; 27:941–949.

15. Wong S, Janssen I Ross R. Abdominal adipose tissue distribution and metabolic risk. Sports Med. 2002; 33:709–726.

16. Wang Y, Rimm EB, Stampfer MJ, Willett WC, Hu FB. Comparison of abdominal adiposity and overall obesity in predicting risk of type 2 diabetes among men. Am J Clin Nutr. 2005; 81:555–563.

17. Kim SK, Kim HJ, Hur KY, Choi SH, Ahn CW, Lim SK, Kim KR, Lee HC, Huh KB, Cha BS. Visceral fat thickness measured by ultrasonography can estimate not only visceral obesity but also risks of cardiovascular and metabolic diseases. Am J Clin Nutr. 2004; 79:593–599.

18. Mohamed-Ali V, Goodrick S, Rawesh A, Katz DR, Miles JM, Yudkin JS, Klein S, Coppack SW. Subcutaneous adipose tissue releases interleukin-6, but not tumor necrosis factor-alpha, in vivo. J Clin Endocrinol Metab. 1997; 82:4196–4200.

19. Fontana L, Eagon JC, Trujillo ME, Scherer PE, Klein S. Visceral fat adipokine secretion is associated with systemic inflammation in obese humans. Diabetes. 2007; 56(4):1010–1013.

20. Humphries SE, Luong LA, Ogg MS, Hawe E, Miller GJ. The interleukin-6-174 G/C promoter polymorphism is associated with risk of coronary heart disease, systolic blood pressure in healthy men. Eur Heart J. 2001; 22:2243–2252.

21. Joffe YT, Collins M, Goedecke JH. The relationship between dietary fatty acids and inflammatory genes on the obese phenotype and serum lipids. Nutrients 2013; 5:1672– 1705.

22. Albani D, Batelli S, Polito L, Prato F, Pesaresi M, Gajo GB, De Angeli S, Zanardo A, Galimberti D, Scarpini E, Gallucci M, Forloni G. Interleukin-6 plasma level increases with age in an Italian elderly population (“The Treviso Longeva”-Trelong-study) with a sex-specific contribution of rs1800795 polymorphism. Age (Dordr). 2009; 31(2):155–162. doi: 10.1007/s11357-009-9092-5. Epub 2009 Apr 18.

23. Rodrigues KF, Pietrani NT, Bosco AA, Campos FMF, Sandrim VC, Gomes KB. IL-6, TNF-α, and IL-10 Levels/Polymorphisms and Their Association with Type 2 Diabetes Mellitus and Obesity in Brazilian Individuals. Arch. Endocrinol. Metab. 2017; 61:438– 446. doi: 10.1590/2359-3997000000254.

24. Popko K, Gorska E, Demkow U. Influence of Interleukin-6 and G174C Polymorphism in IL-6 Gene on Obesity and Energy Balance. Eur. J. Med. Res. 2010; 15(Supple 2):123–127. doi: 10.1186/2047-783X-15-S2-123.

25. Jurečeková J, Drobková H, Šarlinová M, Babušíková E, Sivoňová MK, Matáková T, Kliment J, Halašová E. The Role of Interleukin-6 Polymorphism (Rs1800795) in Prostate Cancer Development and Progression. Anticancer. Res. 2018; 38:3663–3667. doi: 10.21873/anticanres.12643.

26. Wernstedt I, Eriksson AL, Berndtsson A, Hoffstedt J, Skrtic S, Hedner T, Hultén LM, Wiklund O, Ohlsson C, Jansson JO. A common polymorphism in the interleukin-6 gene promoter is associated with overweight. Int J Obes Relat Metab Disord. 2004; 28(10):1272–1279. doi: 10.1038/sj.ijo.0802763.

